# Early Prediction of Hemodynamic Shock in the Intensive Care Units with Deep Learning on Thermal Videos

**DOI:** 10.1101/2020.11.12.20230441

**Authors:** Vanshika Vats, Aditya Nagori, Pradeep Singh, Raman Dutt, Harsh Bandhey, Mahika Wason, Rakesh Lodha, Tavpritesh Sethi

**Affiliations:** Indraprastha Institute of Information Technology, Delhi-110020, India; CSIR-Institute of Genomics and Integrative Biology, New Delhi-110007, India; Academy of Scientific and Innovative Research (AcSIR), Ghaziabad, 201002, India; Shiv Nadar University, Tehsil Dadri, Greater Noida, Uttar Pradesh-201314; All India Institute of Medical Sciences, Department of Pediatrics, New Delhi-110029, India

## Abstract

Shock is one of the major killers in Intensive Care Units and early interventions can potentially reverse it. In this study, we advance a non-contact thermal imaging modality to continuous monitoring of hemodynamic shock working on 103,936 frames from 406 videos recorded longitudinally upon 22 patients. Deep learning was used to preprocess and extract the Center-to-Peripheral Difference (CPD) in temperature values from the videos. This time-series data along with heart rate was finally analyzed using Long-Short Term Memory models to predict the shock status up to the next 6 hours. Our models achieved the best area under the receiver operating characteristics curve of 0.81 ± 0.06 and area under the precision-recall curve of 0.78 ± 0.05 at 5 hours, providing sufficient time to stabilize the patient. Our approach, thus, provides a reliable shock prediction using an automated decision pipeline, that can provide better care and save lives.

## Introduction

Hemodynamic shock is an acute and initially reversible condition that can act as an early signal to future-end organ failure[1]. It manifests due to inadequate blood perfusion than what is essentially required. Such a condition could cause tissue malfunction leading to rapid organ deterioration which can eventually result in death. The mortality rate due to delay in detection and treatment of such shock is as high as 34% in the ICU patients admitted in developing countries[2]. Thus, proactive monitoring and therapy of hemodynamic shock can prevent the risk of impending organ failure and mortality[3],[4],[5]. However, a delayed or insufficient monitoring can increase the risk of hemodynamic shock. Modern ICUs are equipped with a large number of sensors that generate humongous amounts of data besides a large number of clinical investigations. ICU clinicians and nursing staff are exposed to this large quantity of data on a real-time basis. The overall burnout can go as high as 80.5% which can contribute to delayed or insufficient care[6]. Moreover, the resource-limiting areas have a higher risk of shock ignorance mainly due to a lack of technological advancement and a low doctor-to-patient ratio[7]. Most of the methods dealing with the shock in current times are invasive or require repeated contact, making the patient prone to hospital-acquired infections. The non-invasive ways such as Non-Invasive Blood Pressure monitoring and Ultrasonography are not continuous but require contact with the delicate skin of infants which are infection-prone too[8]. Thus, the use of artificial intelligence (AI) and frugal open-source technologies such as low-cost non-contact thermal monitoring can be effective for shock management.

We have created a body thermal imaging data cohort for the pediatric population under the SafeICU initiative which warehouses 1.5 million hours of physiological vitals data, laboratory investigation, treatment charts, doctors and nurse investigation charts[9]. We have used these data to build an automated hemodynamic monitoring and prediction of future onset of hemodynamic instability pipeline. The most recent minimally invasive way to get sufficient data is to work on the thermal images[9]. Thermal image data is now increasingly being used in medical literature for breast cancer detection[10], and even diabetes mellitus (type 1) detection[11]. Thermography of the skin surfaces is also being used for preliminary hypertension assessment[12]. The studies have found out that the possibility of shock can be determined using the temperature difference observed between the abdomen and foot of the patient, i.e. Centre-to-Peripheral Difference (CPD)[13],[14]. The feature has been exploited along with some vitals and the use of machine learning methods such as Histogram of Oriented Gradients features with Random Forest classifier for detection and prediction of shock with a single snapshot of an image at a time point[15]. But a single instance might not be able to give robust results because of limited information. Plus, using the handcrafted features for machine learning algorithms might cost us time. To expand upon the work, we aim to reduce the manual tasks performed to calculate the features using deep learning methods and work on the continuous time-series data, improving the efficacy of detection and prediction.

In this work, we developed a future risk prediction model (see Figure 1) using continuously streamed videos of body infrared patterns and state-of-the-art Computer Vision (CV) and Deep-Learning (DL) techniques. We carried out thermal monitoring as a part of the SafeICU warehouse. We constructed an automated CPD extraction pipeline by body part segmentation on thermal videos using deep-learning models. Each time-point was labeled for shock and non-shock using age-specific shock-index[16]. Time-series data of the extracted CPD signals, combined with heart-rate and the labels were pre-processed to case-control cohort design. We constructed cohorts at 0 hour for detection and 1-6 hour lead-time for prediction tasks. We then used multivariate Long Short Term Memory (LSTM)[17] models to detect and predict the future onset of the shock. We finally evaluated the performance metrics of the models using a 10 fold cross-validation method. Thus, the aim of our work was to build an automated, minimal contact pipeline for proactive hemodynamic monitoring. Automated detection & prediction will help prevent the adverse outcomes related to hemodynamic instability. Hence, the impact this problem creates and the scope of extending the solution make this study worthwhile in saving the lives of many.

**Figure 1:**
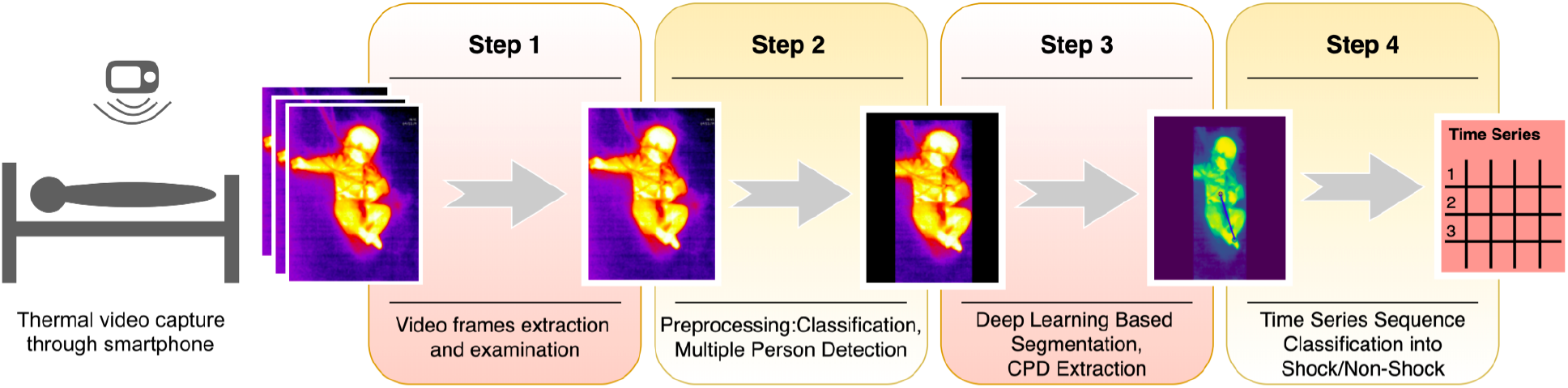
Shock Prediction Steps. The summary of the shock pipeline shows the steps from video frame extraction to shock prediction. Step 1 comprises sampling videos to extract frames. Step 2 classifies frames into covered or uncovered, while also finding the presence of multiple people in the frame and mask them, to avoid confusion. The masked images are then input to the ResUNet based segmentation model and CPD is hence extracted. The series of CPDs are then passed through a time series sequence classifier and finally, the predictions are made for shock for the next 6 hours.

## Results

### Patient Characteristics, pre-processing and cohort building

Statistical Inferences of the cohort characteristics between the shock and non-shock groups are depicted in Table I. Shock and Non-shock conditions were derived using the Age-specific shock index cut-off values. It can be observed that the most significant difference between the two categories is in the heart rate, as expected, along with the respiratory rate. We obtained a median length of stay of 6.808 days with a total of 132 shock instances and 274 non-shock instances. A total of 130 infants and children (male, female), aged between 0.2-204 months, who were admitted and had the arterial line recordings were considered in this study. A total of 76 (of 52 patients) out of 181 thermal videos recorded through Android Smartphone attached Seek Thermal® camera were considered for analysis after screening for quality and concomitant vitals measurements. Out of these, 44 thermal videos from 30 children (male, female) were chosen based on the availability of their abdomen and feet uncovered. 16 out of 44 videos were of a duration of 1 min or less. Since the project looks at long windows of longitudinal data, only 26 long videos (2+ hours; 7200s or more), with their vitals present, remain consisting of 22 unique patients.

**Table 1.** Cohort characteristics and statistical significance of control (non-shock) vs affected (shock) classes. The p-values were calculated using either Wilcoxon rank-sum test (W) or Student’s t-test (t) after testing for normality by the D’Agostino-Pearson normality test. (n - Number of sequences, IQR - Interquartile Range)

| Variable | Non-shock Seq ( n = 274) | Shock Seq (n = 132) | Statistical Tests |
| --- | --- | --- | --- |
|  | Median (IQR) | Median (IQR) | p-value (W/t) |
| Age (months) | 54.44 (59.43) | 75.89 (107.02) | 0.6087 (W) |
| Arterial Systolic Blood pressure, mm Hg | 130.75 (11.95) | 128.09 (25.21) | 0.0014 (t) |
| Systolic Blood pressure, mm Hg | 106.00 (12.00) | 102.00 (5.00) | 0.002 (W) |
| Heart rate, per min | 111.52 (20.27) | 143.17 (63.81) | < **0.0001 (t)** |
| Respiratory rate, per min | 25.85 (9.66) | 22.87 (13.20) | < **0.0001 (W)** |
| O<sub>2</sub> saturation (SpO<sub>2</sub>)% | 97.86 (2.62) | 98.32 (3.45) | 0.9932 (W) |

### Segmentation of abdomen and feet achieved a total dice loss of 0.0391 using ResUNet

Since thermal images lack texture, it is important for the model to recognise the structural shape features. ResUNet was specifically used to make the task possible on relatively less available thermal data, and to capture both the local and global information from the image. A dice loss of 0.0391 (Dice coefficient = 0.9609) with a Binary Cross Entropy loss of 0.0692 was achieved for segmenting out abdomen, feet, and the background. The mode intensities of the segmented areas were then used to find out the Central-to-Peripheral Difference and hence build the longitudinal models from continuous long-duration videos. The results are showcased in Supplementary Table S1.

### The LSTM model was found to be the best performing

We compared three models to finally arrive at the best performing one. Linear-Mixed Effects[18], Random Forest[19], and LSTM[17] were tested at various time points from the observation taken. Based on various metric evaluations, LSTM was found out to be the best performing on our given time series data, and hence was chosen as the primary choice for our study. The F1 score comparison of the three models is shown in Figure 2, and the rest of the evaluations are shown in the Supplementary Fig. 1.

**Figure 2:**
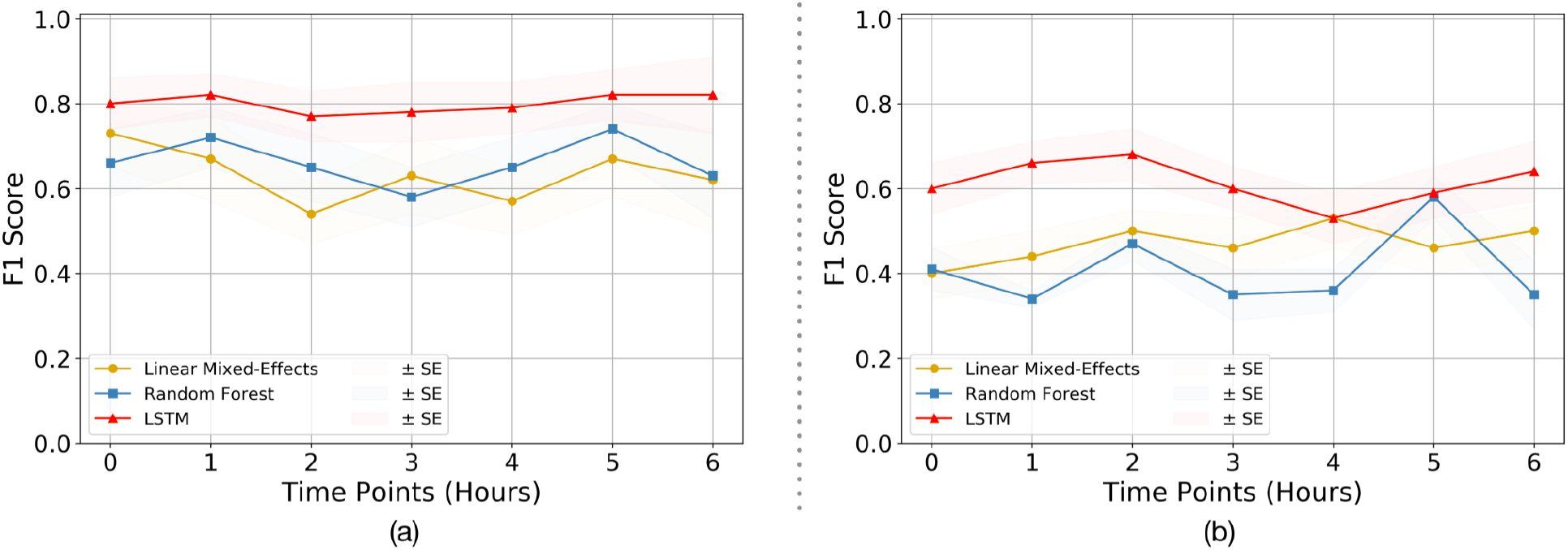
Quality Assessment of Models. The F1 score comparison for the three models (i.e. Linear Mixed-Effects model, Random Forest, and LSTM) tested on (a) CPD & heart rate, and (b) CPD parameter only. It can be observed that the LSTM model outperforms the other two models in both cases, making it the primary choice of this research. The rest of the comparison plots are depicted in the Supplementary Fig. 1.

### Evaluation of the LSTM model at different lead time reveals that CPD increases the model performance

The SAFE-ICU resources allowed us to match the time sequences with their corresponding states of shock/non-shock for the next 6 hours since the observation was taken. We tested the LSTM model performance for 1 to 6 hours of lead time on CPD and heart rate as shown in Table 2. As shown in Figure 3, we got the best performance at AUROC 0.81 ± 0.06 and AUPRC of 0.78 ± 0.05 at 5 hours. This gives a good time window to alert the medical practitioners to start the therapy for preventing the advent of shock.

**Table 2.** Performance of the proposed model predicting the presence of Shock/Non-shock using automated CPD and HR. The Time Pt. column depicts the subsequent hours from the time of taking the observation, at which the results were recorded. The unequal number of shock and non-shock sequences is due to the absence of patient data with the increasing number of hours. (S/NS - Number of Shock/Non-Shock sequences present, AUPRC - Area Under Precision Recall Curve, AUROC - Area Under Receiver Operating Characteristics, PPV - Positive predictive value, NPV - Negative predictive value, D - Detection, P - Prediction)

| Time Pt. | S/NS | AUPRC | AUROC | Accuracy | Sensitivity | Specificity | PPV | NPV | Youden |
| --- | --- | --- | --- | --- | --- | --- | --- | --- | --- |
|  |  | Mean (SE) |  |  |  |  |  |  | Mean |
| 0hr (D) | 132, 274 | 0.79 (0.06) | 0.78 (0.05) | 0.85 (0.03) | 0.74 (0.06) | 0.92 (0.02) | 0.86 (0.05) | 0.84 (0.03) | 0.50 |
| 1hr (P) | 115, 271 | 0.71 (0.06) | 0.76 (0.04) | 0.83 (0.04) | 0.83 (0.04) | 0.82 (0.06) | 0.80 (0.06) | 0.88 (0.02) | 0.42 |
| 2hr (P) | 125, 253 | 0.56 (0.05) | 0.69 (0.04) | 0.83 (0.02) | 0.72 (0.06) | 0.90 (0.03) | 0.82 (0.05) | 0.84 (0.04) | 0.56 |
| 3hr (P) | 133, 232 | 0.67 (0.06) | 0.74 (0.06) | 0.83 (0.04) | 0.69 (0.08) | 0.93 (0.04) | 0.88 (0.06) | 0.82 (0.04) | 0.57 |
| 4hr (P) | 123, 242 | 0.75 (0.05) | 0.75 (0.05) | 0.81 (0.04) | 0.70 (0.06) | 0.94 (0.03) | 0.90 (0.05) | 0.78 (0.06) | 0.64 |
| 5hr (P) | 120, 247 | 0.78 (0.05) | 0.81 (0.06) | 0.84 (0.04) | 0.76 (0.07) | 0.94 (0.02) | 0.88 (0.05) | 0.84 (0.05) | 0.62 |
| 6hr (P) | 124, 228 | 0.66 (0.10) | 0.73 (0.06) | 0.89 (0.03) | 0.82 (0.08) | 0.92 (0.04) | 0.81 (0.09) | 0.95 (0.02) | 0.62 |

**Figure 3:**
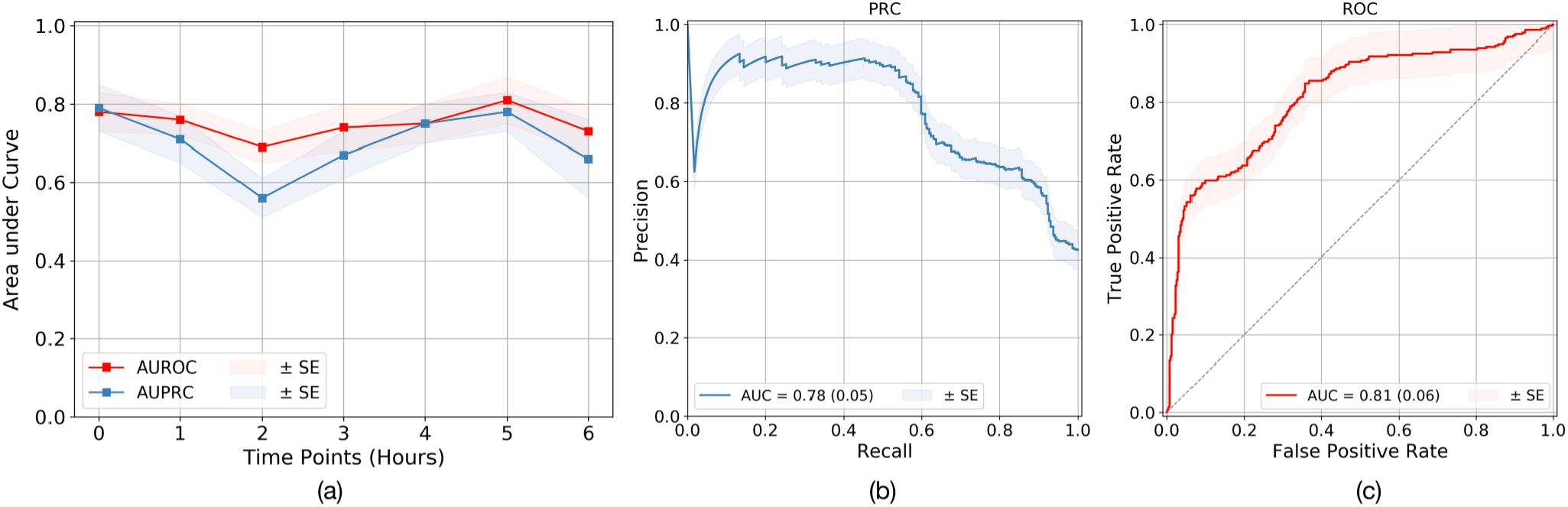
Quality Evaluation on LSTM model. (a) Quality evaluation of the LSTM time series classification models at lead points of up to 6 hours. The best performance of (b) AUPRC and (c) AUROC was obtained at a 5-hour prediction. The rest of the performance metric results are demonstrated in the Supplementary Fig. 2. The results of all are shown in Table 2. The Standard Error (SE) for each is calculated from cross-validation, by taking k=10.

## Discussion

The study presents a computer vision and deep learning based continuous and non-contact shock detection and prediction model which leverages Center-to-Peripheral Difference (CPD) as one of its main parameters. Since hemodynamic shock can lead to organ failure and eventually to death in the ICU, its prediction on time can save lives. Care needs to be taken to evaluate the parameters in a non-invasive way such as not to cause the infections by the contact. Since the field of thermal image inspection has only started to be explored, it can thus be leveraged, along with some non-invasively monitored vital parameters, for a prediction of shock. The use of deep learning methods proves to be really beneficial in reducing manual preprocessing and increasing the accuracy of the methods.

In this study, we have extended our previous work on hemodynamic shock prediction with longitudinal continuous monitoring of body thermal patterns which are previously found to be predictive for future shock prediction[15]. The longitudinal monitoring of temperature gradient opens up a rich source of information about patients’ physiology. The underlying stochastic patterns can have a discriminative value for future hemodynamic shock risk. We leveraged these reasonings to extract the center to peripheral intensity difference from the thermal images in a time-series fashion. We do so by applying our data-specific trained screening filters for covered and uncovered patient detection models, multiple person detection models, which are followed by segmenting the body parts into abdomen and foot using artificial intelligence based models called Res-UNet. The extracted CPD (see Figure 4) time-series along with vitals time-series were used to predict the future (1-6 hr) hemodynamic shock using sequence models called Long-Short Term Memory (LSTM). This model was also compared with Random Forest and Linear Mixed-Effects models, known for working with longitudinal data. It was observed that the LSTM model outperformed the other two. We also trained and checked the performance of models that can classify the length of video and images for future risk of hemodynamic shock. The class of algorithms in deep learning called convolutional neural networks (CNNs) which have shown state-of-the-art performance for image classification was used for the task. However, we found the domain features such as CPD, performed better than the classification based direct image and video classification for future risk of shock. Even after many experiments performed for a direct classification of images using the concepts of TV Chambolle[20] denoising, data augmentation, and undersampling/ oversampling, we were able to get the best AUROC of only 0.60 for shock detection using ResNet-50[21]. This can be because of the cluttered background with diapers and tubes, and an increased region of interest for the information extraction from a limited variety of images. This also tells the importance of the domain-specific features over automated CNN based approaches. We built the models for multiple time-points till 6 hours which showed the best performance at a lead time of 5 hours. The metric AUPRC and F1 score are the most significant for an imbalanced dataset such as ours as it doesn’t get biased by the presence of true negatives and thus gives a clearer perspective of a classifier’s utility. The results till 6 hours show a promising window with a good prediction rate which can prove to be helpful for the doctors to help find a buffer time prior to the shock event and therefore start the treatment.

**Figure 4:**
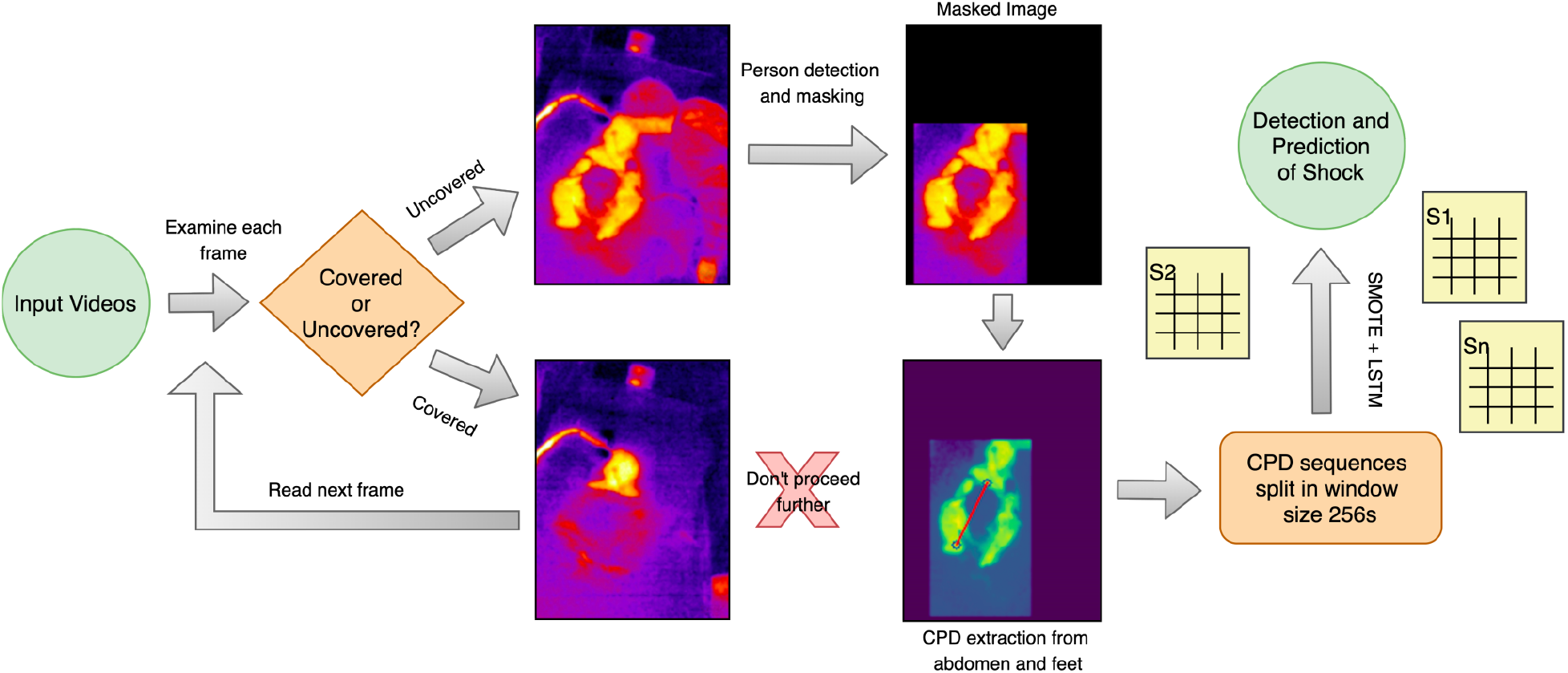
Shock Prediction Pipeline. Illustration of the pipeline followed for the detection and prediction of shock and no-shock. Each frame of the video was examined for uncovered/covered. The uncovered frames were then filtered off the presence of people other than the patient present in the frame. The frame was finally passed from the segmentation and CPD extraction model, further collecting the sequences used for LSTM time series classification. The time sequences had CPD and heart rate as features and an appropriate window length of 256s was chosen. Since the data was highly imbalanced, SMOTE upsampling method was used in training the LSTM model. The detection and prediction of shock were done at 0 hour and for the next 6 hours respectively.

The limitations of our method arise during the quality data procurement. Monitoring thermal patterns could be hard at times when the patient is being covered or being operated on; caregivers can also block the recordings. But, these issues can be resolved by keeping ∼4.3 minutes of uncovered slots for monitoring. Nevertheless, we have built an AI system that is efficient enough to identify the covered and uncovered images, and can remove the caregiver or person other than the patient. Thus, our system is able to extract the thermal patterns efficiently. Even though using CPD as a co-acting parameter eliminates the use of invasive arterial monitoring and cuff-based blood pressure monitors which can cause infections due to the repeated use on multiple patients. Our models only require as minimal as ∼4.3 minutes of thermal recordings which do not recognise or identify an individual’s identity, hence is safe for patient privacy and does not take much time away from the time assigned for care. Our methods can be extended beyond the intensive care settings, i.e. to help in the community based monitoring by the developing countries’ rural healthcare workers, such as the Accredited Social Health Activist (ASHA), run by the women promoting accessible healthcare in rural Indian settings. This can be made possible by training them to use a specifically designed frugal mobile app for thermal shock detection and prediction, which can work with readily available Android/iOS smartphones, with only model weights preinstalled. The results can be made more robust by expanding the dataset by including more number of patients in the study. Also the dataset can be made varied and generalised by including observations from multiple other clinical sites. Nevertheless, the study performed illustrates a great non-invasive and minimal feature architecture that promises to be a life-saver by informing the clinicians about shock well in advance.

## Methods

### Data collection and study design

- **SafeICU Framework:** All the data reported in this research was collected from the Paediatric ICU of AIIMS, New Delhi, a tertiary care hospital in India. There were 8 beds including neonatal beds in the Paediatric ICU. We set up our servers to collect the real-time physiological vitals periodic data. We also warehoused laboratory investigations, daily doctors’, nurses’ notes, treatment charts, and thermal imaging.
- **Ethical Approval and Patient Consent:** The study was carried out with the approval of the Pediatric Intensive Care Unit of All India Institute of Medical Sciences, New Delhi, India. Since thermal images only capture infrared radiation, these don’t reveal patient identity and the study did not involve any contact or change in routine patient care. Hence a waiver of consent was sought and granted by the Institute Ethics Committee (Ref. No. IEC/NP-211/08.05.2015, AA-2/09.02.2017). All experiments were performed in accordance with relevant guidelines and regulations as approved by the ethics committee.
- **Vital records from multi-parameter monitoring:** For monitoring the patients, Paediatric ICUs are equipped with Mindray TM monitors. We installed a dedicated server for querying and storing the vitals streaming data from Central Monitoring Station (CMS). Client socket programming with respect to the device protocols was used. In-house software was written for Health Level 7 (HL7) Standards[22] based querying of the CMS. These vital data were received at the resolution of 15 seconds for unsolicited data and 1 second for the real-time data. To receive streaming data, 64*1024 bytes of character array was used as a buffer. Pipe delimited text file was generated every day at 00:00 hour. Software code was written in such a way that it automatically logs the data into a text file on a daily basis. These high-resolution vital data have been warehoused for all the ICU patients starting from February 2016 to May 2020.
- **Cohort based on Binary shock index:** The SAFE-ICU described earlier has warehoused over 1.5 million patient-hours of monitoring data from the PICU. It is used to extract time-stamped vital data for the patients at 0-6 hour heart rate and blood pressure recordings. Shock index was calculated as the ratio of median heart rate and median non-invasive blood pressure or the arterial systolic blood pressure. This was calculated over the median of moving sequential windows of 30 data points at a resolution of 15s. Shock-Index Paediatric Age-Adjusted (SIPA) is used to compute shock/no-shock age-specific binarized outcome for each patient [16].
- **Thermal Imaging:** Standard thermal video capturing and operating procedures were followed, in order to ensure minimal disturbances by the extraneous factors, say, patient positioning, device handling etc. (Supplementary Methods S1). Thermal cameras only capture infrared radiation, so as to make sure that study does not reveal the patient’s identity. The camera was placed properly and at a good distance from the patient so that there was no direct contact involved nor any change in patient routine care. The thermal videos were captured in a standard color-scale guaranteeing that the full body of infants was visible. Thermal videos of every single patient were collected through an Android Smartphone attached Seek Thermal® camera at different time points on different days. Thus each patient possibly has different values for shock-status, which in turn eliminates bias due to the patient’s propensity characteristics, say gender, age, etc. Vital data with respect to the time-stamp of the videos were extracted from the data warehouse at 15s intervals (SAFE ICU)[9]. A comparison was made between the shock and non-shock groups using either Wilcoxon rank-sum test or two-tailed Student’s t-test, after testing for normality by D’Agostino-Pearson normality test using GraphPad Prism version 6.00, GraphPad Software, La Jolla California USA, www.graphpad.com.

### Classification into Covered and Uncovered

The patients in ICU are kept under observation for a long duration. Since it is a very critical area, the patients are kept covered by a blanket most of the time. The blankets are removed for a short period of time generally only when a nurse or a doctor comes to provide the care. To train the data, images were augmented and normalized by their mean and variance especially extracted out to suit the thermal data. A ResNet-152 architecture was trained using PyTorch[23] framework in Python3[24] to classify each frame into covered and uncovered, i.e. abdomen and feet are visible. The model was finally implemented on videos sampled at 1fps.

### Multiple Person Detection

In the intensive care unit, caregivers tend to provide care to the patient. The caregiver might come into the field of the camera mounted over the bed. For the CPD extraction task, there is a need to filter out the presence of this additional person, so as not to confuse the algorithm between the caregiver and the patient. A variety of images of the patient alone and along with the person/caretaker was taken and augmented. The frames could just have been discarded but a few videos in the dataset contained the presence of the caretaker throughout the duration for which they were captured. The now visible area could be further used for CPD extraction. The thermal images were manually annotated for which they were captured. The now visible area could be further used for CPD extraction. The thermal images were training the multiple person detector.

We used YOLOv3[25] in PyTorch having DarkNet53 as its base architecture. Finally, the trained detector was evaluated for the IoU(Intersection over Union) area; the best performing detector model was used for detecting and masking the caregiver.

### Segmentation and CPD Extraction from Abdomen and Feet

Nagori A. et al.[15] proved that the probability of shock depends directly on Center-to-Peripheral Difference (CPD). For this study, the abdomen has been taken as the center and the peripheral is taken to be the foot. The images were annotated manually and pixel-wise using js-annotator-tool. The target maps contained 3 one-hot encoded layers corresponding to the abdomen, feet, and background. The input images were normalized with the mean and variance especially extracted from the distribution of the dataset in use. Appropriate image padding was done to ensure the aspect ratio of the images remains the same in case of any change in the input dimension. To account for a low dataset of pixel-wise segmented images for training, a ResUNet with ResNet-18, pre-trained on ImageNet, was used as an encoder. UNet[26], being specifically introduced to segment the less abundantly found medical data, helps to gather more local and global information even in the dearth of data, and thus efficiently segmenting out the images. The skip connections from the encoder to the decoder helps the model to keep the original pixels at that particular scale in consideration while recreating at the decoder and thus learning finer details efficiently. Smaller skip connections in ResNet-18 encoder helps to deal with the problem of vanishing gradients and thus make the learning more efficient[23]. A cutoff threshold was set on the predicted outputs to remove any weakly predicted pixels. The area detected was used as a region of interest in the original image and the mode of the detected probabilities was taken as the point of temperature extraction from the segmented out abdomen and feet. The difference was divided by the abdomen value to keep CPD robust from the thermal noise.

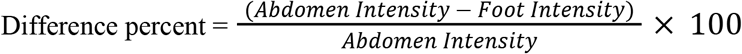

### LSTM Time Series Sequence Classification

The videos were sampled at 1 fps to extract the CPD data from every uncovered window possible. Windows of 256 data points corresponding to 256s (4.26 min, padded, if necessary) were taken as an input to the LSTM based classifier. The windows less than 256 are padded with 0s and the windows greater than 256 are split in an overlapping fashion, when necessary. Each CPD, along with the heart rate at its corresponding time point was taken to finally label it with the shock index, and hence the presence of shock/non-shock. The missing heart rate data at certain points was imputed with linear interpolation if the missing data was less than 10% of the time series length. Since the data is highly imbalanced with more non-shock sequences, the training data is augmented with the SMOTE[27] oversampling method. The LSTM sequence classifier was followed by a series of dense layers with a dropout of 0.2, which then passed through a sigmoid layer to output the binary shock index, and hence, the occurrence of shock/no-shock.

### Linear Mixed-Effects and Random Forest sequence classification on tsfresh features

The tsfresh[28] features were extracted from 256-length sequences and trained on the same train and validation distributions as the previous LSTM model. ‘Boruta’[29] package from R-language was used for this purpose. Variation Inflation Factor (VIF) is used to reduce multicollinearity in data. If the VIF value exceeds 10, then the collinearity is considered problematic and hence that particular variable causing it should be removed. The remaining features were used to train the linear mixed-effects and random forest models.

### Direct Classification of thermal images/videos for future risk of shock

Apart from CPD extraction, an attempt was made to classify into shock/no-shock by directly giving the whole images/videos as the input. In one direction, we tried to classify each video frame read at a time and conducted experiments with several modern architectures based on convolutional neural networks (CNN). The concepts of TV Chambolle denoising, data augmentation and undersampling/oversampling, were used to get the best shock detection AUROC of 0.60 using ResNet-50. Also, the information extracted from a single image frame can be very limited. So instead, we tried to use direct and continuous video samples of length 256s as an input to a conjunction of various CNN and LSTM models, trained in a time distributed manner. Being a fundamental extension of the direct image classification problem, it suffered from similar limitations.

### Outcome variable - Binary shock index

The SAFE-ICU initiative has enabled this research to gather the PICU data and extract the vitals and the corresponding time stamps at the 0th hour (time of video capturing) and at the next 6 hours. Shock index was taken as the median heart rate and median non-invasive blood pressure or the arterial systolic blood pressure, for moving sequential windows of 30 data points at a resolution of 15s. Shock-index Paediatric Age-Adjusted (SIPA) was then used to compute the age-specific binary outcome for each patient.

### Time Points

The time at which the video was captured was taken as the 0th hour, and the predictions of shock/no-shock were performed for the next 6 hours.

### Model Evaluation

The video data was first partitioned patient-wise such as to keep train, validation, and test sets unseen from each other. For the 10-fold cross-validation, the data was partitioned with the ratio of 60:20:20 into these three sets in a stratified manner, i.e. keeping the distribution of low-percentage shock class comparable in all three sets. The training data was augmented for the low-found shock class using SMOTE oversampling method; the validation and test sets remain unchanged in their size in each respective fold. The model analysis was mostly done on the Area Under Precision-Recall Curve (AUPRC) and Area under Receiver Operating Characteristic (AUROC) curve. Other standard metrics like F1-score, PPV, NPV, Specificity, and Sensitivity, were evaluated at the Youden’s Index (J)[30]. Since there is a high significance of prevalence in the medical domain, calculating the metrics at Youden’s Index becomes important.

## Data Availability

Data-sets used in the study are available from the corresponding author on reasonable requests.

## Data Availability

The data is available on a reasonable request from the corresponding author.

## Acknowledgements

This work was supported by the Wellcome Trust/DBT India Alliance Fellowship IA/CPHE/14/1/501504 awarded to Tavpritesh Sethi. Tavpritesh Sethi and Pradeep Singh also acknowledge the support received from the Department of Science and Technology vide project DST/INT/ISR/P-21/2017. We also thank Mr. Varun Prakash and Mr. Anil Sharma for the technical support provided at PICU, AIIMS, New Delhi.

## Author contributions

Concept & Design: TS, RL, AN, PS. Standard thermal video data acquisition: PS, TS, RL. Vital data acquisition: AN, PS, TS, RL. Cohort construction using vital data and thermal videos: PS AN. Segmentation Models & Extraction of CPD: VV. Multiple person detection & covered-uncovered detection: VV. LSTM, Models & Evaluation: VV, AN, HB. Mixed-effects and Random forest models & Evaluation: PS, AN, VV. Statistical analysis: VV AN. Direct classification models: RD, HB, MW. Interpretation of Results: VV, AN, PS, TS. Manuscript & Revision: VV, AN, PS, RL, TS. The final version of the manuscript was approved by all the authors involved in this research.

