## Supplementary Material for "Early Prediction of Hemodynamic Shock in the Intensive Care Units with Deep Learning on Thermal Videos"

### Classification into covered or uncovered patients

Since the patients are kept under observation for long hours in intensive care conditions, it is possible for them to remain wrapped in a blanket for most of the time. Using ResNet-152, we were able to classify the video frames into covered/uncovered to get a hold of any window during which the blanket is removed. An accuracy of 95.52% was achieved, with an F1 score of 0.9589 (Precision=0.9722, Recall=0.9459), creating fewer chances of false positives.

### Multiple person detection using YOLOv3

The patient is barely left alone when he/she is uncovered under long hours of observations. To remove any chance of confusion between the temperatures and the body parts that the following segmentation algorithm might face, YOLOv3 was deployed for multiple person detection which achieved an IoU of 0.71206. The other person and the background were then masked to keep the patient in focus for CPD extraction.

**Supplementary Table S1.** Performance of models in the preprocessing steps in the tasks of classification, object detection and segmentation

| Classification | | | |
|---|---|---|---|
| ResNet-152 | | | |
| Precision | Recall | F1-Score | Accuracy |
| 0.9722 | 0.9459 | 0.9589 | 0.9552 |

| Object Detection | Segmentation | | |
|---|---|---|---|
| YOLOv3 | ResUNet | | |
| IoU | BCE Loss | Dice Loss | Total Loss |
| 0.7121 | 0.0692 | 0.0391 | 0.0542 |

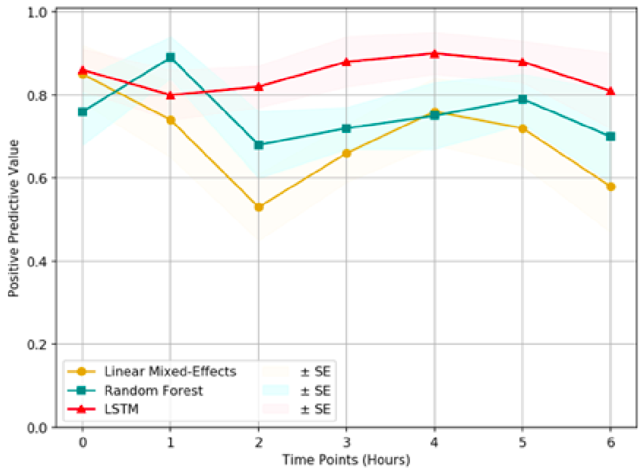


(a) PPV Plot

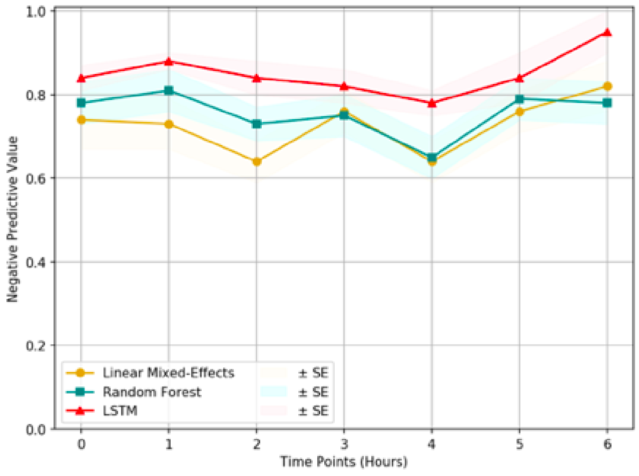


(b) NPV Plot

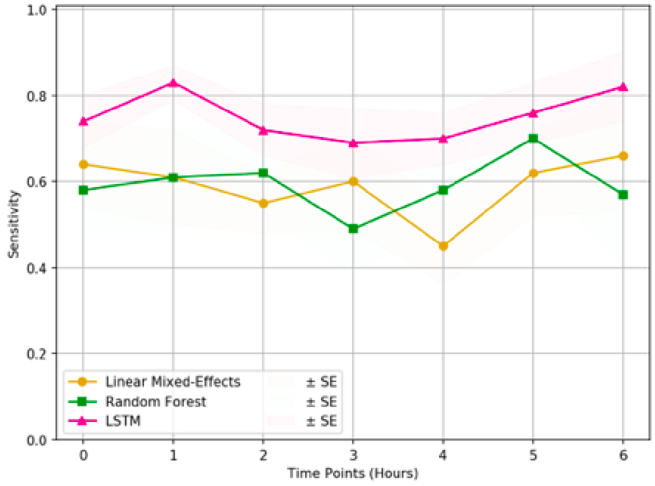


(c) Sensitivity Plot

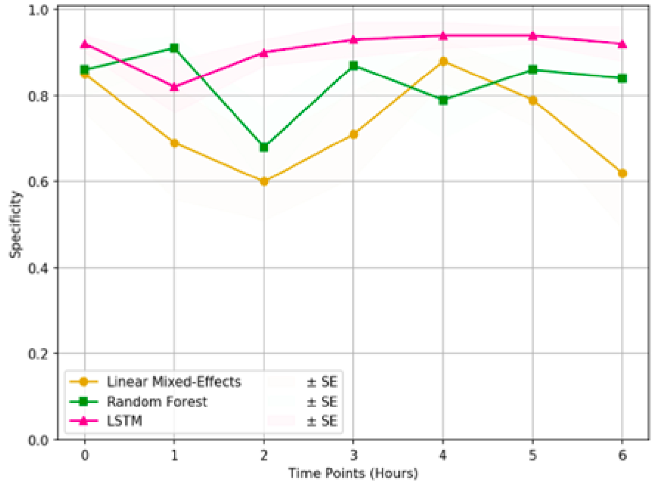


(d) Specificity Plot

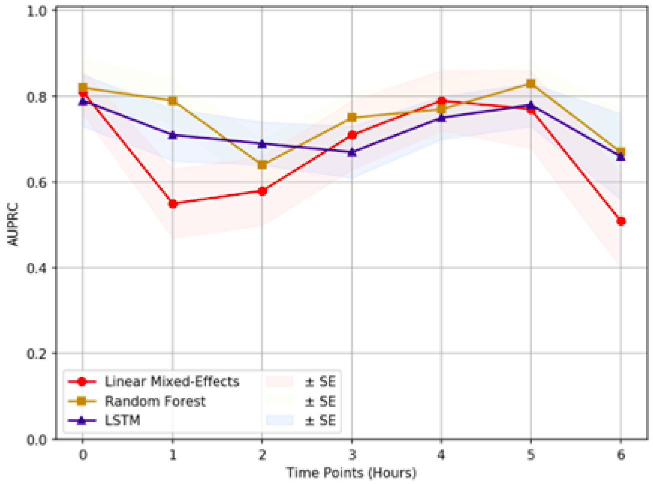


(e) AUPRC plot

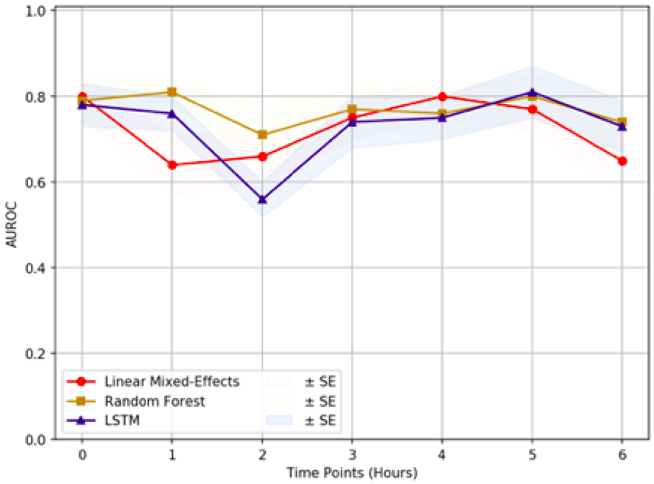


(f) AUROC plot

**Supplementary Figure 1. Comparison of classification models.** The three models were compared based on their ability to classify the sequence data. It can be observed that the LSTM model outperforms the other two with respect to the **(a)** Positive Predictive Value, **(b)** Negative Predictive Value, **(c)** Sensitivity and **(d)** Specificity. **(e)** Area Under Precision-Recall Curve (AUPRC) and **(f)** Area Under Receiver Operating Characteristics (AUROC) of the models are comparable. Since our research data exhibits a high data imbalance, F1 score and AUPRC metrics are the primary considerations. LSTM model displays surpassing F1 score along with PPV, NPV, sensitivity, and specificity, making it the prime choice for this study.

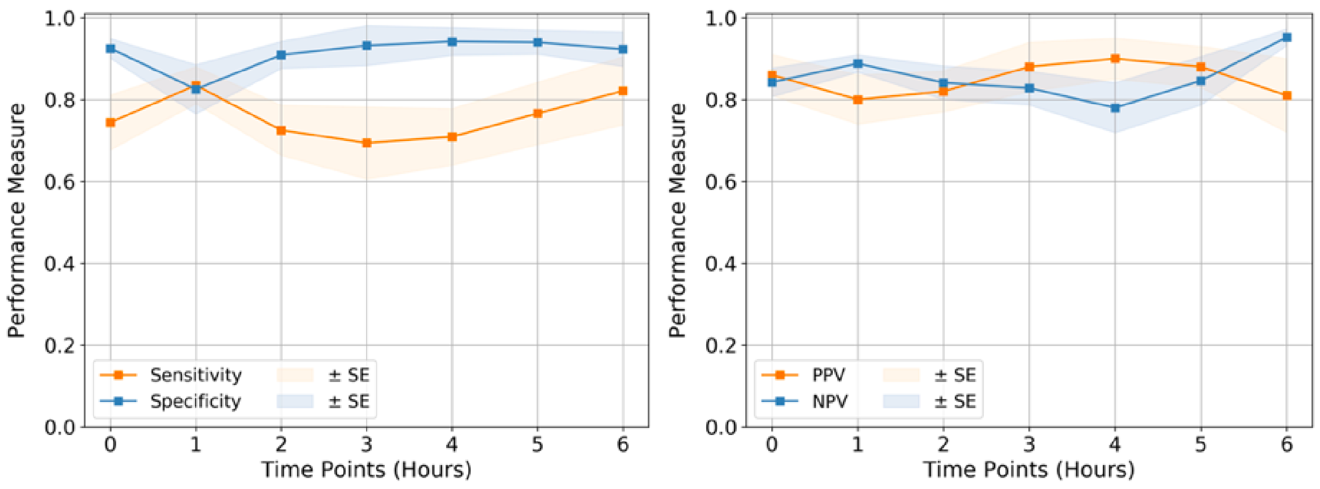


(a) Sensitivity and Specificity Plot

(b) PPV and NPV plot

**Supplementary Figure 2. Quality Assessment:** Comparison of **(a)** Sensitivity and Specificity and **(b)** Positive Predicted Value (PPV) and Negative Predicted Value (NPV) by LSTM model at each time point till 6 hours.

#### Shock detection at 0 hour using LSTM

The CPD was extracted from every uncovered window possible using the segmented out abdomen and feet regions in a continuous way with videos sampled at 1fps. Keeping in consideration the Unique Health IDs of patients and then performing SMOTE upsampling, the observation windows of 256s (i.e. 4.26 min) were passed into LSTM networks, along with heart rate as an additional covariate, for sequence classification. 10-fold stratified cross-validation gave a mean AUPRC of 0.796 and mean AUROC of 0.788.

### Supplementary Methods S1

#### Standard operating protocol (SOP) for capturing thermal videos

1. Before entering the ICU, make sure that one has to wear shoe covers, a scrub, and a disposable gown.
2. Need to connect the Seek Thermal Camera to the available Smartphone and then open the Seek Thermal app.
3. Ensure that the Seek Thermal app is set to a high-resolution setting and to an "Iron Theme" before capturing videos. Wall mount hanger is used to hold a smartphone. Seek Thermal Camera is placed at a good distance from the bed and at a certain height in order to make sure that patients on the bed are not disturbed.
4. For each patient recorded in this session, the following steps were followed:
   - Making the patient comfortable is the most important part. The camera was placed properly and at a good distance from the patient so that there was no direct contact involved nor any change in patient routine care.
   - In the presence of a nurse, a smartphone is hanged using a wall mount hanger.
   - MSExcel sheets are used for data collection. Data such as UHID, bed number, date, and time of the patients on this session were recorded for identification purposes.
   - After capturing videos remove the mobile hanger safely and save the data on a server, daily basis, into an identified folder with the patient's UHID, bed information, and date.

**Supplementary Table S2.** The metrics observed for direct classification of images for shock detection. (SGD - Stochastic Gradient Descent, AUROC - Area Under Receiver Operating Characteristics)

| Model | Optimizer | AUROC | Accuracy |
|---|---|---|---|
| ResNet-50 | SGD | 0.60 | 65% |

**Supplementary Table S3.** Statistical significance analysis using the Hanley and McNeil formula[1]. It is specifically used as our models are stratified k-fold cross-validated, and the train and test sets would be independent within each fold, but might not be across the folds. Stratified cross-validation violates the IID condition and hence Wilcoxon/student's t-test cannot be used. Hanley, et. al. also state that single-tailed tests can be used in such cases, and a value near to the cutoff p-value can be considered as evidence that this observation may not be completely random. Hence, single-tailed analysis was performed with an alpha cutoff value of 0.10. It can be observed that AUPRC, our primary metric in use for data imbalance conditions, displays its statistical significance for most of the time points for the time window under our consideration.

| Time Pt. | AUPRC | | AUROC | |
|---|---|---|---|---|
| | p-value | Significant? | p-value | Significant? |
| 0hr | 0.116102 | ~Yes | 0.406949 | No |
| 1hr | 0.174508 | ~Yes | 0.169814 | ~Yes |
| 2hr | 0.002146 | Yes | 0.006782 | Yes |
| 3hr | 0.407027 | No | 0.418395 | No |
| 4hr | 0.077194 | Yes | 0.412780 | No |
| 5hr | 0.215846 | No | 0.144732 | ~Yes |
| 6hr | 0.000095 | Yes | 0.014258 | Yes |
